# A modified SEIR model to predict the COVID-19 outbreak in Spain and Italy: simulating control scenarios and multi-scale epidemics

**DOI:** 10.1101/2020.03.27.20045005

**Authors:** Leonardo López, Xavier Rodó

## Abstract

After the spread of SARS-CoV-2 epidemic out of China, evolution in the pandemic worldwide shows dramatic differences among countries. In Europe, the situation of Italy first and later Spain has generated great concern, and despite other countries show better prospects, large uncertainties yet remain on the future evolution and the efficacy of containment, mitigation or attack strategies. Here we applied a modified *SEIR* compartmental model accounting for the spread of infection during the latent period, in which we also incorporate effects of varying proportions of containment. We fit data to quarantined populations in order to account for the uncertainties in case reporting and study the scenario projections for the 17 individual regions (CCAA). Results indicate that with data for March 23, the epidemics follows an evolution similar to the isolation of 1, 5 percent of the population and if there were no effects of intervention actions it might reach a maximum over 1.4*M* infected around *April*27. The effect on the epidemics of the ongoing partial confinement measures is yet unknown (an update of results with data until March 31st is included), but increasing the isolation around ten times more could drastically reduce the peak to over 100*k* cases by early April, while each day of delay in taking this hard containment scenario represents an 90 percent increase of the infected population at the peak. Dynamics at the sub aggregated levels of CCAA show epidemics at the different levels of progression with the most worrying situation in Madrid an Catalonia. Increasing alpha values up to 10 times, in addition to a drastic reduction in clinical cases, would also more than halve the number of deaths. Updates for March 31st simulations indicate a substantial reduction in burden is underway. A similar approach conducted for Italy pre- and post-interventions also begins to suggest substantial reduction in both infected and deaths has been achieved, showing the efficacy of drastic social distancing interventions.

## 1 Introduction

Epidemiological reports for the current situation of COVID-19 out of China indicate Europe is one of the centers of the pandemic, and that nearly 200 countries/regions worldwide have reported cases[1]. Particularly in Italy and Spain a large outbreak of SARS-CoV-2 infection is underway. Given the lack of previous exposure to this virus, future predictions of both the levels of virus spread in the global naive population and the acquired herd immunity cannot be properly anticipated. Data on the last similar pandemic one century ago remains limited for obvious reasons, despite aftermath reports exist [2]. Policymakers in the most affected countries face a difficult situation when trying to balance between draconian public health actions and keeping economy alive, as impact on health of severe economic measures is also well-known [3]. The fact that the 1918 Spanish flu pandemic had a second deadly epidemic wave, presumably caused by mutations in the *H*1*N* 1 viral strain [4], has also stimulated a vivid debate on whether actions to take now should take into account this uncertain future. Under this scenario, optimal action on the COVID-19 pandemic is hard to fathom. And more so, for the extent of pre-symptomatic and asymptomatic infections is not narrowly constrained (e.g. up to 86 percent[5]). In China, regions as well as local governments, including Hubei, tightened preventive measures to curb the spreading of COVID-19 since Jan. 2020 [6]. Many cities in Hubei province were locked down and many measures implemented, such as tracing close contacts, quarantining infected cases, promoting social consensus on self-protection (e.g. wearing face mask in public area, minimum social distances). However, in other areas, the extent and efficacy of the so-called confinement or self isolation is doubtful, and facebook data on mobile location and movement showed yet massive people displacements under semi-confinement measures. At a time when success of large-scale social distancing interventions is critical, access to accurate information to ascertain mobility is lacking [7]. Similarly, credible serology tests that could show whether someone has had the infection and recovered are not yet massively deployed, thereby assessment of SARS-CoV-2 prevalence in the population is not established [8].

We followed the approach of Peng et al. [6] and implemented a modified SEIR model that accounts for the spread of infection during the latent period. This novel capacity to significantly spread during the latent period is a distinctive feature of the current SARS-CoV-2 epidemic if compared to SARS in 2003. Therefore, many classical models such as SIR[9, 10, 11], SEIR [11] and SEIJR [12] are not appropriate to describe the outbreak of SARS-CoV-2 in China and elsewhere. Thanks to new data on the average latent period and average time of treatment, this time delay process can be successfully incorporated in a novel dynamical system framework to describe this quite unique dynamics. It has been discussed that at times of ongoing epidemics, and due to errors and under reporting, accumulated numbers of diagnosed cases and even number of deaths might reflect more reliably the extent of epidemic progression than the daily reported new cases. Alternately, and as in Chen 2020 [9] and Peng et al. [6] we also employ the accumulated numbers in time as variables, as it was done also for modelling MERS-CoV in the recent past [13]. Isolating key segments of populations (e.g. vulnerable populations, workers providing essential services, or a territory with too fast growing incidences) to protect them from an uncontrolled epidemic progression, is under vivid discussion as the real extent of viral spread in the population is not well known. Both the limited diagnostic capacity and the lack of clear strategies and mechanisms to quarantine infectious individuals, stand as two of the main limitations to control disease spread. Countries such as South Korea [14, 15] and Taiwan [16, 17] rapidly deployed aggressive contact tracing and quarantine systems [18], understanding that early deployment of resources to try to control initial seeding foci, often not only balances public health criteria but also economics. Given the differences in public health strategies and the varying capacity of the national health systems in each country to tackle the extent of the infection in the population, the growing proportions of undiagnosed infected that eventually show up, are seen to exert an elevated stress on the already saturated health’s system capacities. It is therefore relevant to accurately evaluate the effects of the social distancing actions imposed by governments, such as the individual or territory isolation and interruption of labour activity and/or intra and inter-urban transportation. Such strong provisions of isolation for suspected cases and infection due to contact with undiagnosed individuals are also taken into account in our modelling approach. This same SEIR model has been utilised to compare the effects of lock-down of Hubei province on the transmission dynamics in Wuhan and Beijing [6]. The manuscript is organised as follows: in Section 2, we describe the dynamic model and datasets used to simulate the evolution of the outbreak of SARS-CoV-2 in Spain and in its 17 administrative regions (CCAA), as well as in Italy. The effect on the epidemic curves of the efficacy of the different intervention measures aggregated for Spain, is presented in Section 3.1. Section 3.2 provides estimates of the future number of diagnosed people, fatalities and recovered individuals for each individual CCAA prediction as of March 23 and the main active foci in Section 3.3. Section 3.4 addresses model uncertainties and limitations of our study, and we add results for Italy pre- and post-interventions in Section 3.5. We finally discuss efficacy of the actions in each case and territory in terms of health burden and report and provide an update of the situation up to March 31st.

## 2 Model and Data description

We used a generalized *SEIR* modelling framework similar to Peng et al. [6], that enables for the testing of control interventions. Compared with statistical methods mathematical modelling based on dynamical equations can often provide essential information of the epidemic dynamics. This is particularly true when basic epidemiological parameters are unknown or largely uncertain and more mechanistic understanding is needed, such as for the current COVID-19 disease.

The population is assumed constant due to the rapid disease spread, i.e. the births and natural death have the same value. The recovery rate and death rate are time-dependant. The model also assumes that susceptible individuals (*S*) are contagious upon coming into contact with infected (*I*) individuals not detected. It is assumed, however, that the infected are all quarantined (*Q*) and that they do not have contact with susceptible individuals. In turn, the susceptible population can be protected by confinement by moving to the protected population compartment (*C*). This assumption on *Q* means that hospital infections are not considered under this framework, therefore resulting in a potential underestimation of the real contagion extent. However, we chose this option in an attempt to be conservative and our results should be interpreted as the current best-case scenario. It is assumed also that the protected population does not have contact with the infected individuals and therefore cannot be infected. The COVID-19 dynamics is modelled by the following equations system:

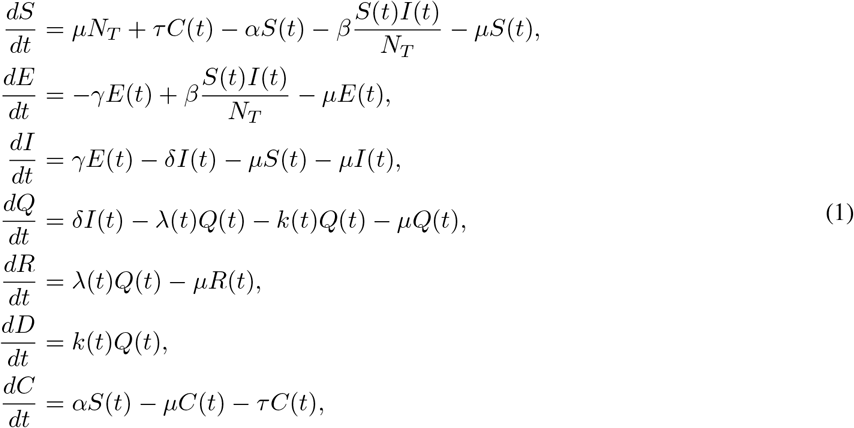

where,S(t) is the susceptible population, C(t) is the confined susceptible population, E(t) is the exposed population, I(t) is the infectious population, Q(t) is the population under Quarantine (infected reported cases), R(t) is the recovered population and D(t) is the dead population by the virus.

The main parameters of the model are the protection rate (*α*), the infection rate (*β*), the incubation rate (*γ*), the quarantine rate (*δ*), the natural death and birth rate(*µ*) (1*/*(80 *\** 365)), the recovery rate (*λ*(*t*)), the mortality rate by the virus (*k*(*t*)) and finally *τ* is the length of the proteciton by confinament (1*/*30) The *α* parameters represent the rate of people being totally protected from infected populations at time *t* and it is used to model the different actions of control of the epidemic by isolation of healthy population.

The parameters *λ*(*t*) and *k*(*t*) are time dependent and for simplicity are modelled as shown next:

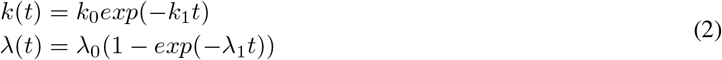

where *k*_0_, *k*_1_, *λ*_0_ and *λ*_1_ are the fitted coefficients [19]. The data for fitting the model to Spain was obtained from public data sources [20, 21] and correspond to the reported cases, recovery cases and deaths. Data is available for the entire country as well as for individual CCAA. Data for Italy is retrieved from the World Health Organization [22].

The model was fitted in a non linear approach by calculating the normalized least-squares error of the model approximation and the infected reported cases. The optimization algorithm used was trust-region-reflective with a maximum number of iterations of 500 and a step size of 6.4*e −* 7. The error for the model fitted for all Spain was 3*e* − 3±0.6*e* − 4. The fitted parameters are summarized in Table 1

**Table 1:** Sample table title

| Comunity | $\alpha$ | $\beta$ | $\gamma$ | $\delta$ | $\lambda_0$ | $\lambda_1$ | $k_0$ | $k_1$ |
| --- | --- | --- | --- | --- | --- | --- | --- | --- |
| Andalucía | 0,015 | 1,073 | 0,776 | 0,610 | 0,062 | 0,017 | 0,171 | 0,970 |
| Aragón | 0,000 | 1,118 | 0,262 | 0,458 | 0,013 | 0,007 | 0,014 | 0,064 |
| Asturias | 0,001 | 1,438 | 0,264 | 0,623 | 0,023 | 0,076 | 0,017 | 1,709 |
| Illes Balears | 0.025 | 1.022 | 0.256 | 0.101 | 0.042 | 0.019 | 0.020 | 0.082 |
| Canarias | 0,007 | 1,257 | 0,182 | 0,335 | 0,035 | 0,030 | 0,005 | 0,002 |
| Cantabria | 0.016 | 1.086 | 0.217 | 0.010 | 0.014 | 0.607 | 0.004 | 0.012 |
| Castilla-La Mancha | 0,000 | 1,339 | 0,571 | 0,750 | 0,027 | 0,044 | 0,015 | 0,000 |
| Castilla y León | 0,015 | 1,124 | 0,575 | 0,467 | 0,059 | 0,027 | 0,045 | 0,219 |
| Catalunya | 0,015 | 1,359 | 1,138 | 0,760 | 0,159 | 0,006 | 0,651 | 0,859 |
| C. Valenciana | 0,015 | 1,155 | 0,862 | 0,674 | 0,090 | 0,007 | 0,447 | 1,267 |
| Extremadura | 0.025 | 1.017 | 0.272 | 0.203 | 0.026 | 0.013 | 0.028 | 0.080 |
| Galicia | 0.021 | 1.835 | 0.915 | 0.892 | 0.076 | 0.004 | 0.003 | 0.000 |
| Madrid | 0,015 | 1,227 | 0,939 | 0,613 | 0,274 | 0,011 | 0,016 | 0,000 |
| Murcia | 0.025 | 1.871 | 0.908 | 1.090 | 0.019 | 0.006 | 0.001 | 0.037 |
| Navarra | 0,002 | 1,012 | 0,681 | 0,686 | 0,048 | 0,002 | 0,003 | 0,000 |
| Euskadi | 0,000 | 1,081 | 0,564 | 0,626 | 0,137 | 0,010 | 0,015 | 0,048 |
| La Rioja | 0,000 | 1,745 | 0,241 | 1,269 | 0,016 | 0,029 | 0,003 | 0,005 |
| Spain | 0,015 | 1,288 | 0,870 | 0,585 | 0,057 | 0,026 | 0,070 | 0,144 |

## 3 Results

### 3.1 Epidemic curves for Spain and the 17 Autonomous Provinces (CCAA)

The model fitting to data aggregated for Spain on reported infected population up to March 23 yields a good approximation to the exponential curve, as well as to the reported evolution in deaths and recovered 1. The model was fitted from the day 1 of the epidemic at the end of February because the initial protection rate is assumed to be 0.

Progression of infected, deaths and recovered track very approximately observations. With data up to March 22, the model predicts the peak in cases around the end of April or early days in May with an error of the approximation of less than 5%. Current scenario may already include to some extent the effects of the partial ‘confinement’ measures imposed on March 13, therefore it is likely that total projected infections would be much higher should these restrictions not exist. More than 1.6million people would have been recovered from SARS-CoV-2 infection by mid May and total deaths for the entire country at that time would rise over 100 thousand people 5.

### 3.2 Evolution of SARS-CoV-2 in the 17 autonomous regions (CCAA)

To approach the degree of colonisation of the virus at smaller levels of aggregation than the entire country, we applied the same model framework to each of the 17 individual *CCAA*. The former was an attempt to model the epidemics spatially and approach scale-derived differences with country aggregated data (e.g. municipality-level data is not yet available). Results are shown in Figure 2 and display varying developmental stages of the COVID-19 epidemic outbreak, thereby recovering the different demographic topologies in the variety of CCAA and Figure 3 shows the extended fitting in order to have a better understanding of different development stages at community level.

**Figure 1:**
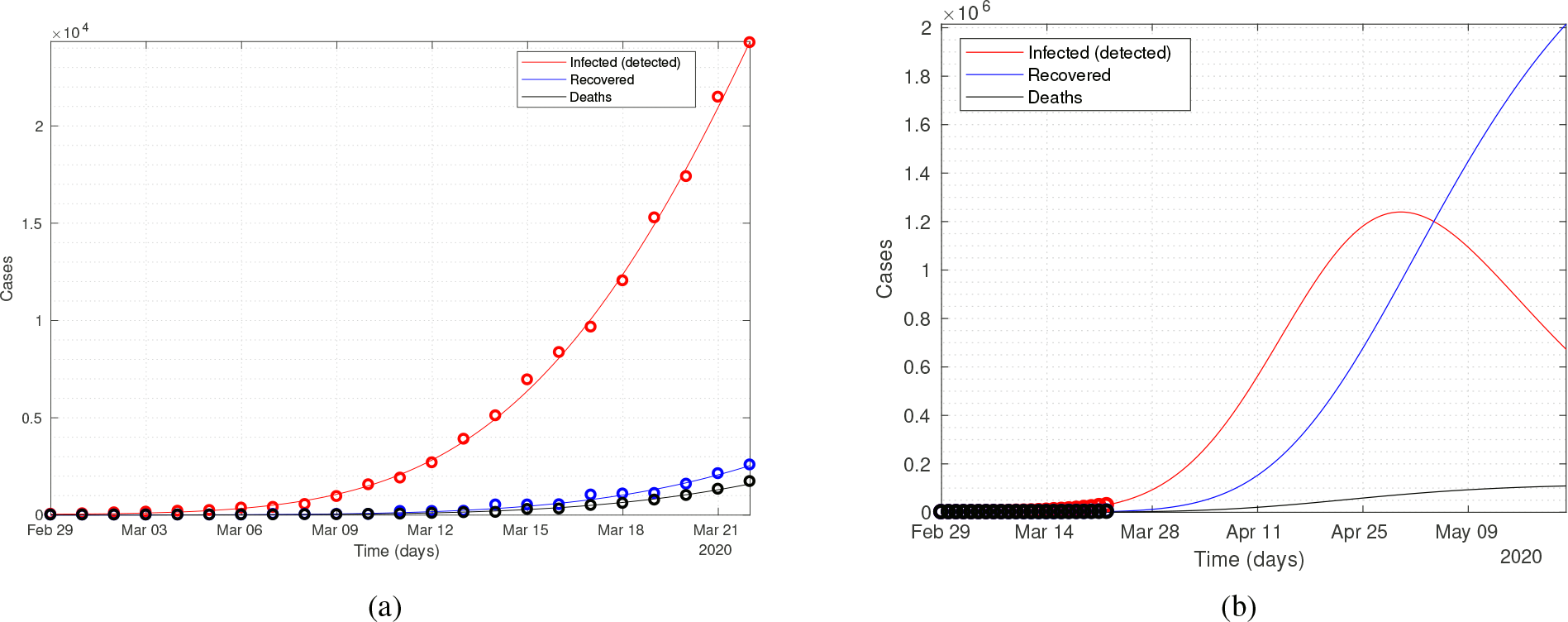
Fitting of the population model implemented on a global scale (all Spain)

**Figure 2:**
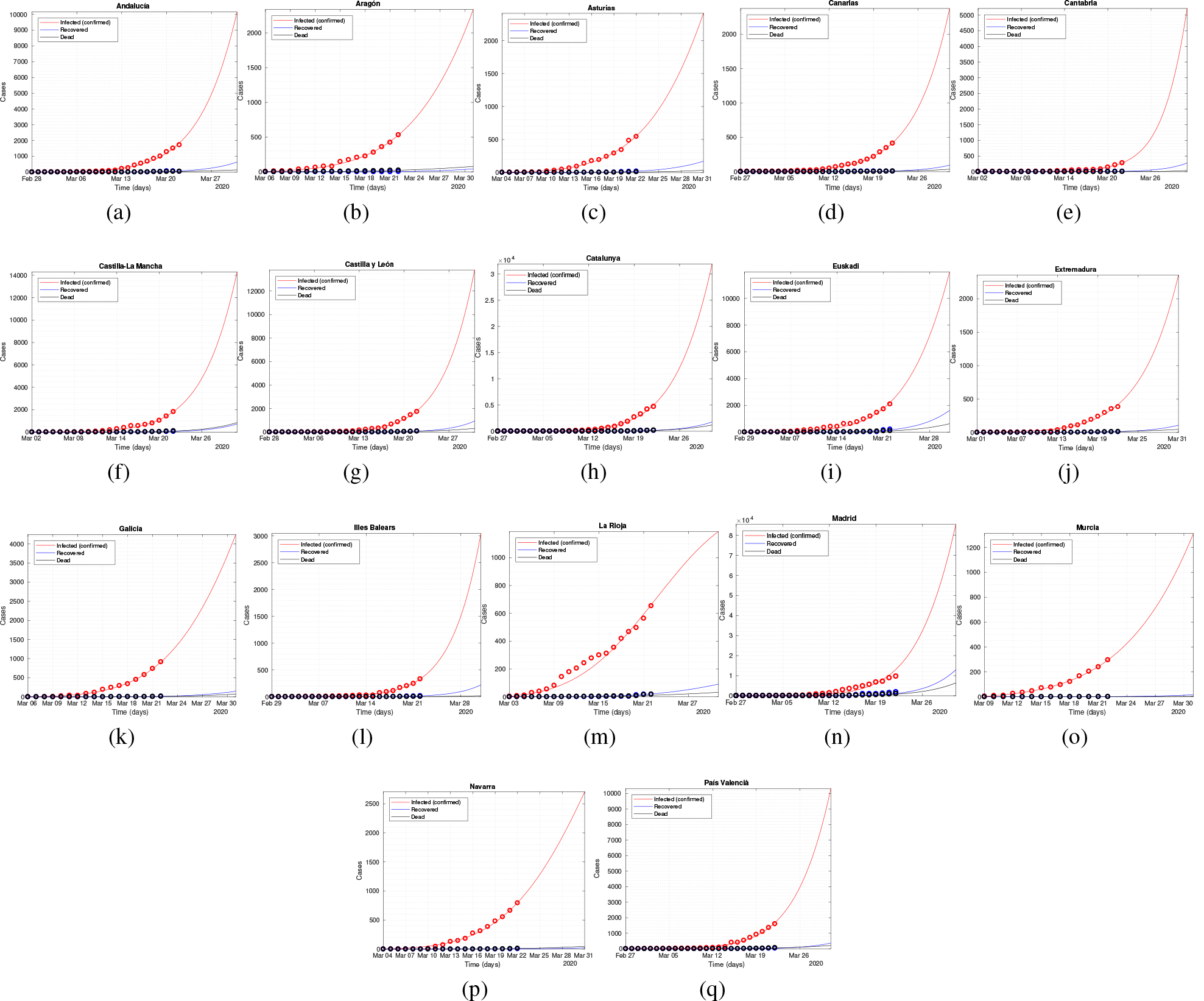
Model fitting for the 17 CCAA of Spain.

**Figure 3:**
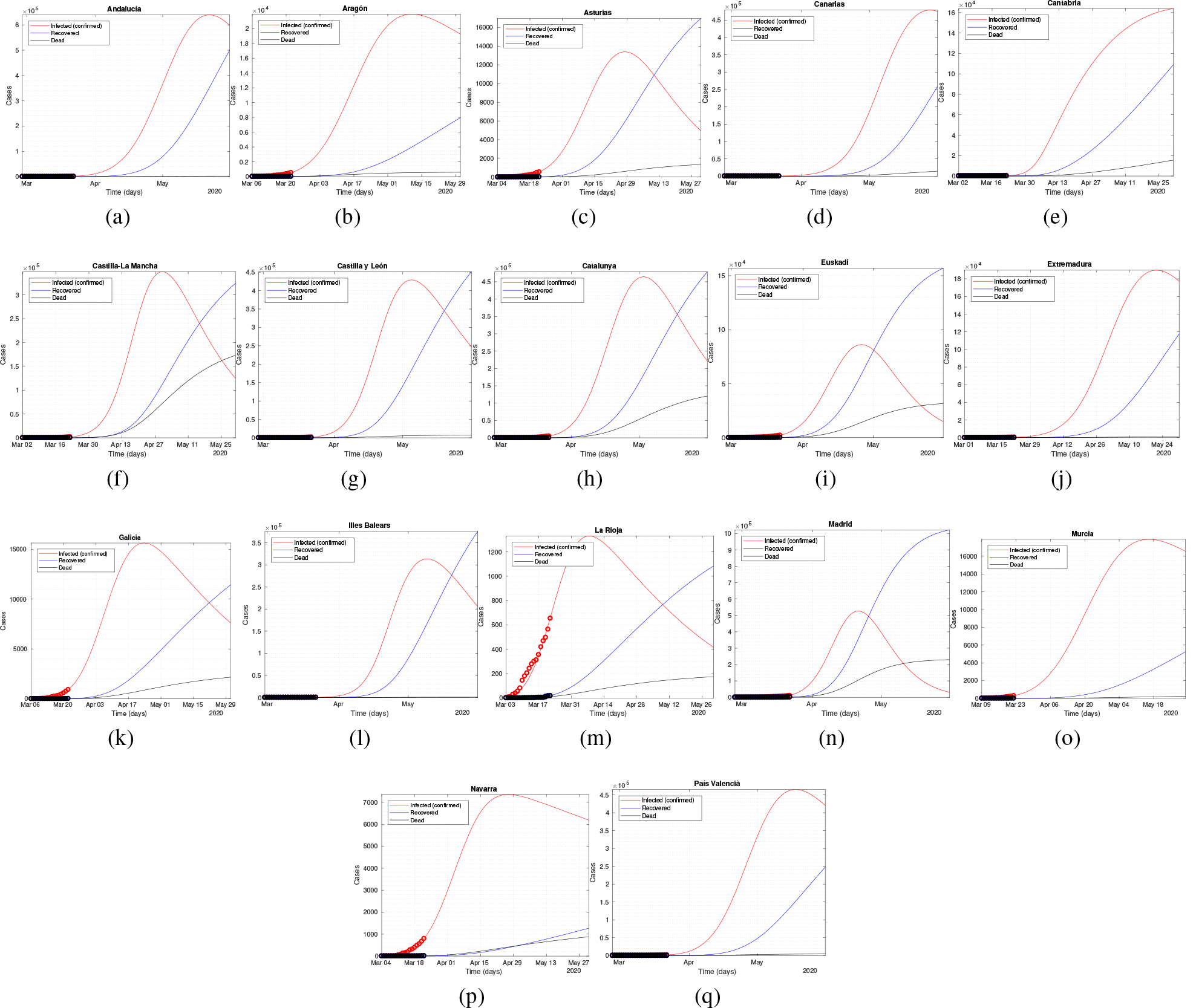
Model extended fitting for the 17 CCAA of Spain.

Largest increases of infected –in absolute terms- are expected according to the CCAA curves, in the coming weeks for Madrid, Catalunya, Castilla La Mancha, Paìs Valencià, Castilla León and Andalucía. All the former display -under the current scenario evolution-peak cases occurring from late April to early May.

To compare Figure 1above with the effect of community dis-aggregation on the evolution of reported cases for Spain, simulations were performed individually and results aggregated together as shown in Figure 4. Results are comparable despite they show relatively larger values for total infected than those arising from the simulations on the country’s total cases.

**Figure 4:**
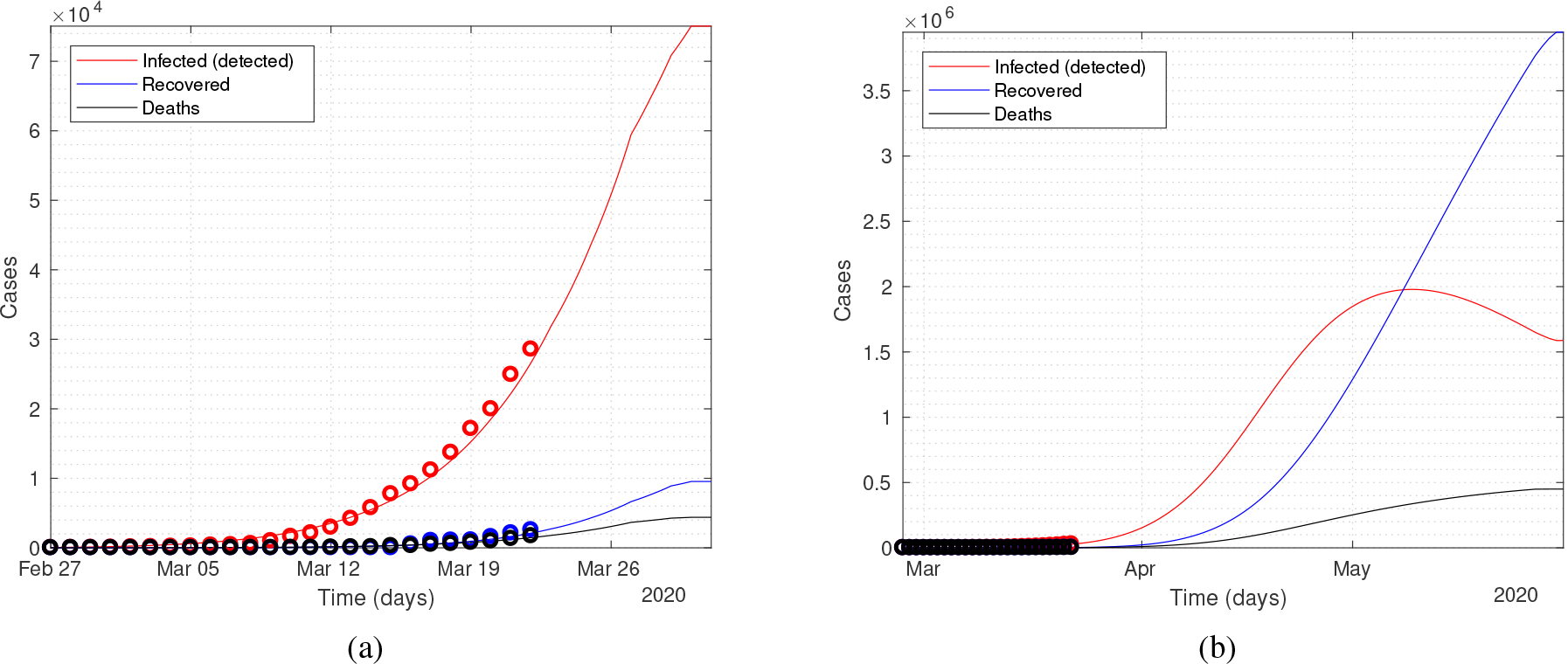
Aggregated CCAA model fitting for Spain.

#### 3.2.1 Control scenarios for Spain

The situation in Spain is at this time yet very severe, based on public data until March 25. To this end, we tested the potential effects of intervention strategies to control and reduce social contacts. Effects of the adopted policies on March, 14 are not yet known or they can barely begin to show up as substantial changes in the epidemic curve at this date (see Appendix for an update to March 31. To help in the evaluation, we generated different scenarios with varying values of *α*, in order to simulate effects of social protection. Increases in *α* were structured in scenarios representing up to 10 times larger values than in that obtained from observations up to March 23. The results of these scenarios are summarised in Figure 5. A drastic decrease in the resulting number of cases can be seen, if we increase the actual daily protection rate around ten times.

**Figure 5:**
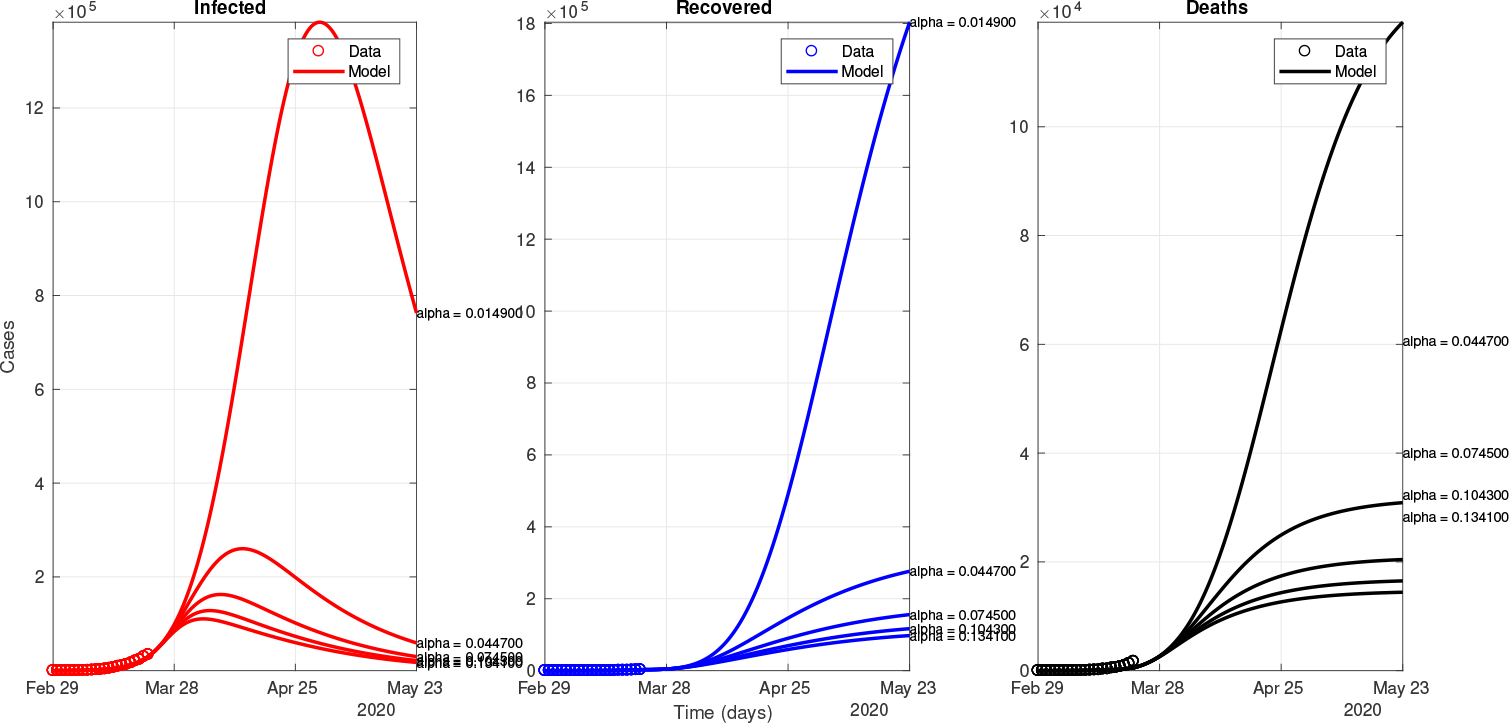
Control scenarios for the global model (all Spain)

Results with the current epidemic evolution for Spain, show that over 1, 400, 000 quarantined infections would be reported by the end of April, with an estimated burden of over 100, 000 deaths. A four times increase in the levels of protected population would represent a huge gain in the number of both clinically diagnosed infections and casualties (Figure 5). A decrease to around 200, 000 infected is achieved and a reduction to below 50, 000 deaths. Much larger reductions in both infected and deaths can be achieved if much stricter measures result in more drastic reductions in the number of effective contacts. Best case scenario simulated with ten times the efficacy of the observed alpha on March, 23, can result in a more recent arrival to a peak of around 100, 000 cases (around early April) and with a decrease in casualties to around 30, 000.

### 3.3 Evolution of the main active foci (Barcelona and Madrid)

Active discussion on the convenience of imposing stricter measures is underway on March 22, with the overall effect of yet acting on the more active COVID-19 foci being under large debate. Given the large associated uncertainties and enormous public health and economic costs, timely actions at this point might represent large gains or losses. To aid in this respect, we simulated the effect of imposing stricter measures of social distancing also on two of the most active foci, namely Madrid and Catalunya. Madrid was the CCAA were the initial outbreak evolved more rapidly and on March 23, it concentrates over 30 percent of total cases and a large majority of deaths. Similarly, Catalunya is now showing a significant increase in new infected and both areas share similar populations albeit Madrid has in comparison, higher population density. Results are shown in Figure 6 for Madrid and in Figure 7 for Catalunya, respectively. Similar conclusions on the large effect of imposing stricter epidemic containment measures can be seen in both regions, even if imposed at the current stage of the epidemic development.

**Figure 6:**
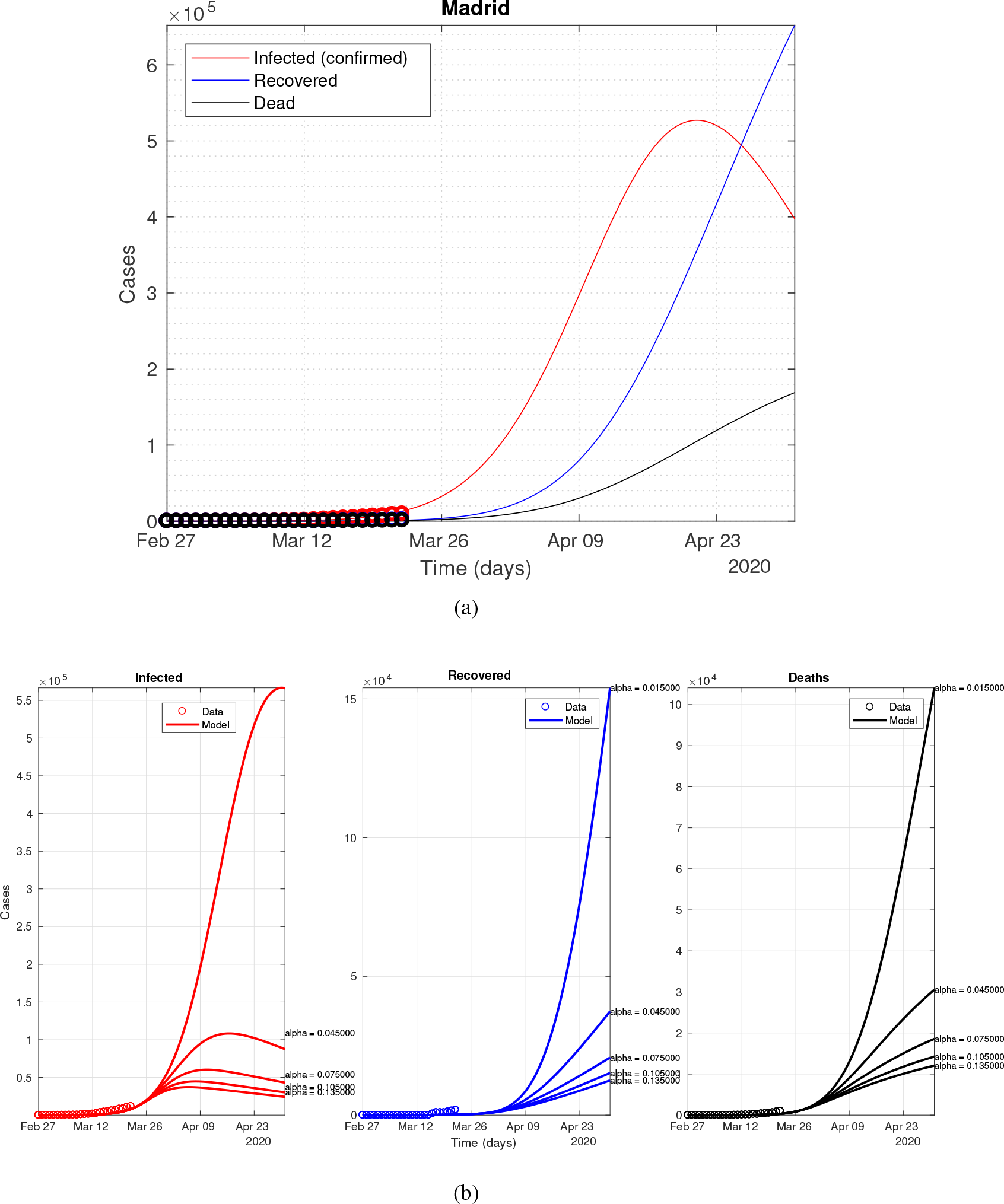
Extended fitting for Madrid.

**Figure 7:**
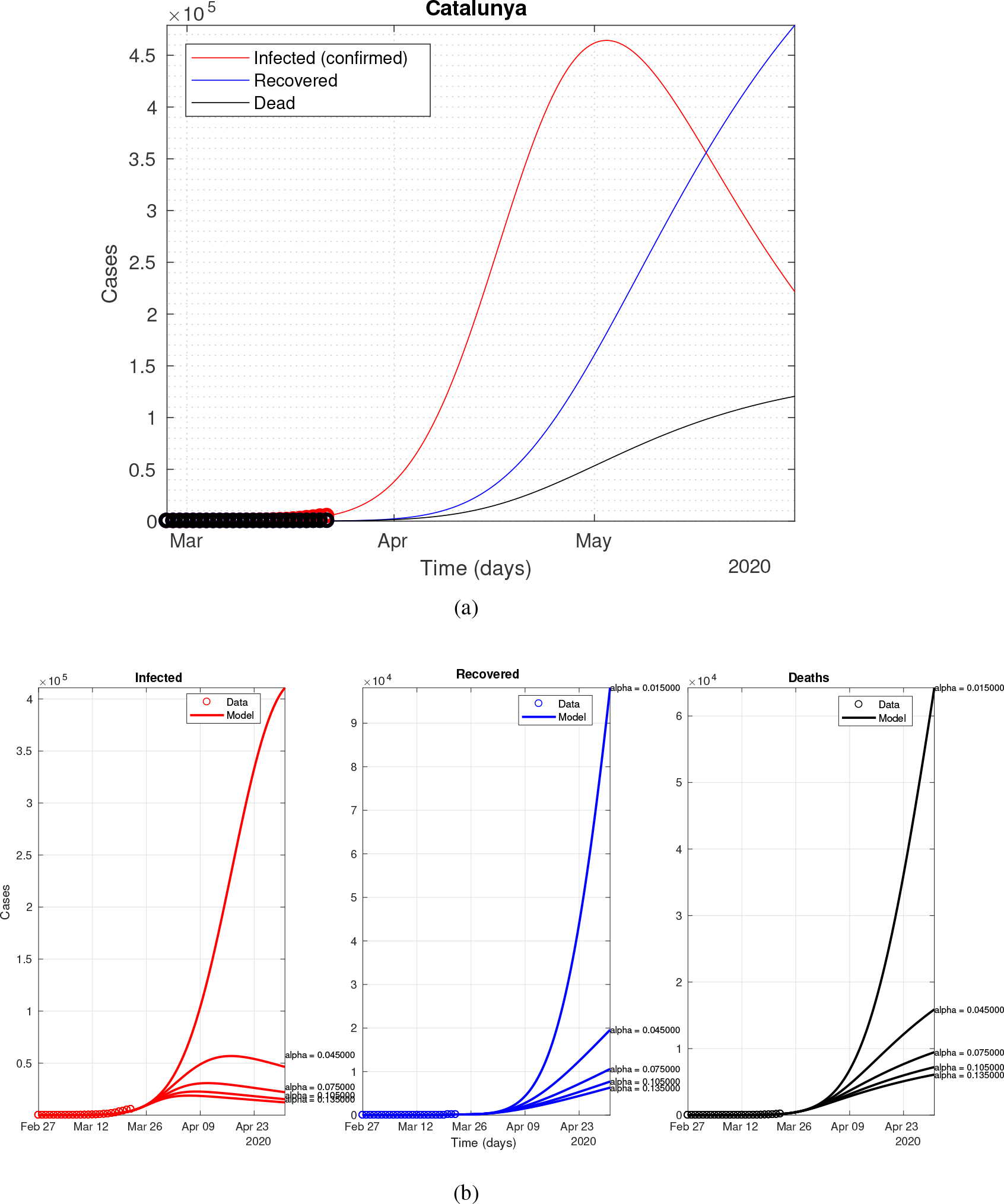
Extended fitting for Catalunya.

### 3.4 Model uncertainties

Because the strength of this type of model, both to adjust and to predict future dynamics, depends on the accuracy in case reporting, it is important to analyze the uncertainties in the adjustment and evaluate the degree of variation of the parameter estimates. To get a better idea on its performance, the model was fitted with successively longer data series, adding a new day to the data window with which the fitting was computed. The shortest time series used was of 5 days from February 29 to March 3 and the longest covered up to March 31. The temporal span of the time series used can seriously affect the value of the fitted parameters. This is especially true at the beginning of the epidemic when the relationship between infected, dead and recovered is not entirely clear. For example if the number of confirmed cases is small, it is difficult to ascertain whether the quarantine has been rigorously applied or not. In addition, such under-representation would suggest that the number of infectious people is much larger than the number of confirmed cases. Results of the different dynamics and the parameter space can be seen in Figure 8. Those parameters most affected are the ones directly related with the infection rate and the recovery rate, as well as the protection rate *α*.

**Figure 8:**
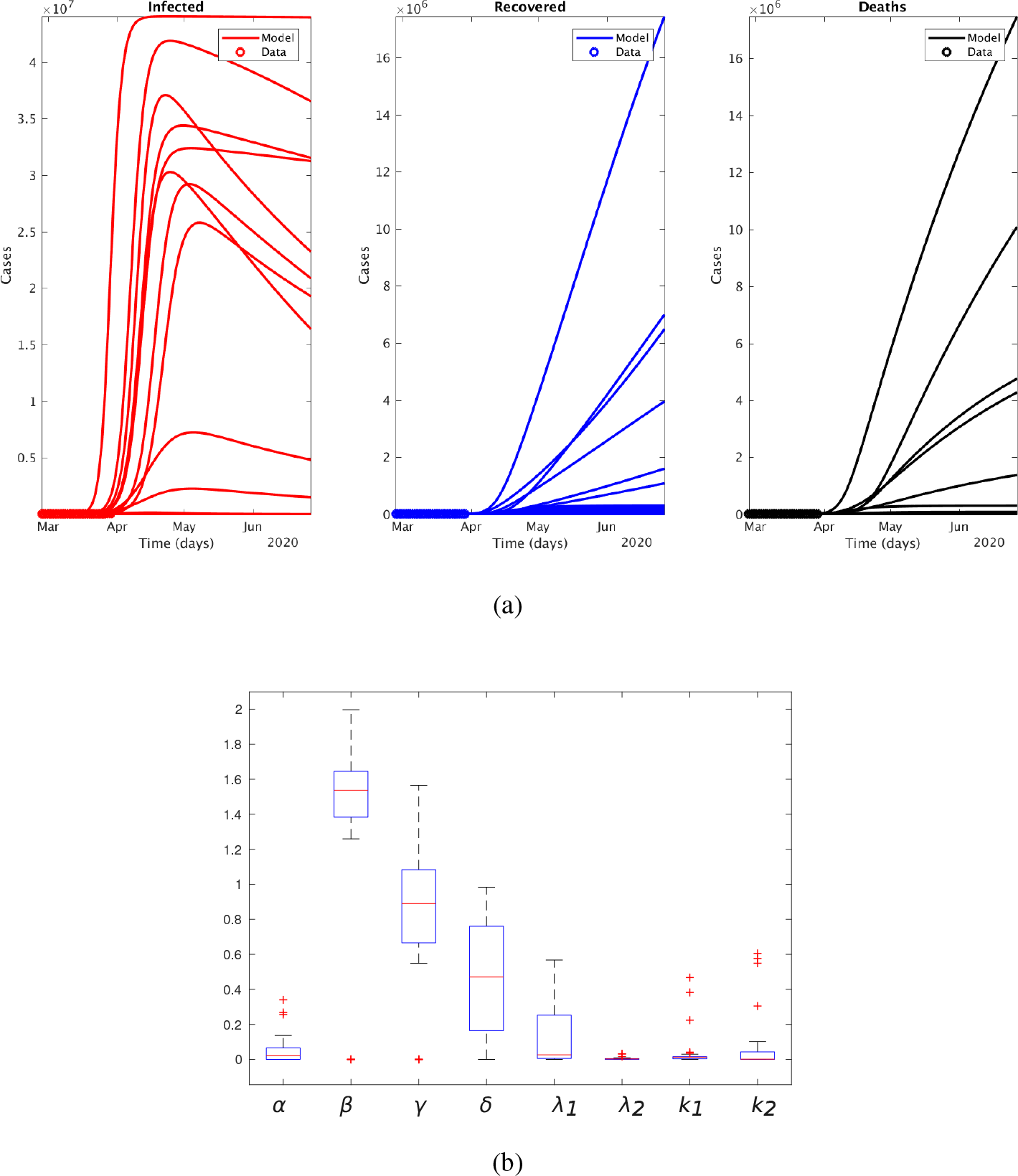
Model uncertainties (a) dynamics obtained by adding new values to the time series and (b) the fitted parameters variation.

### 3.5 The case of Italy

In order to see the performance of applying the same model framework in the other European country with more quarantine cases to date, we fitted the model to the COVID-19 epidemic in Italy. This way, as the situation in Italy precedes that of Spain by around one week, evaluation of control scenarios can be done at a more advanced stage of confinement measures. As the fist cases in Italy were reported in the end of February, two different sets of fittings were performed in order to compare the confinement restrictions imposed by the Italian authorities on March 10*th*. The NMSE in this case was 2.7*e* − 3±3.6*e* − 4 (*ci* = 95%) and the parameters for each case are summarized in Table II.

**Table 2:**
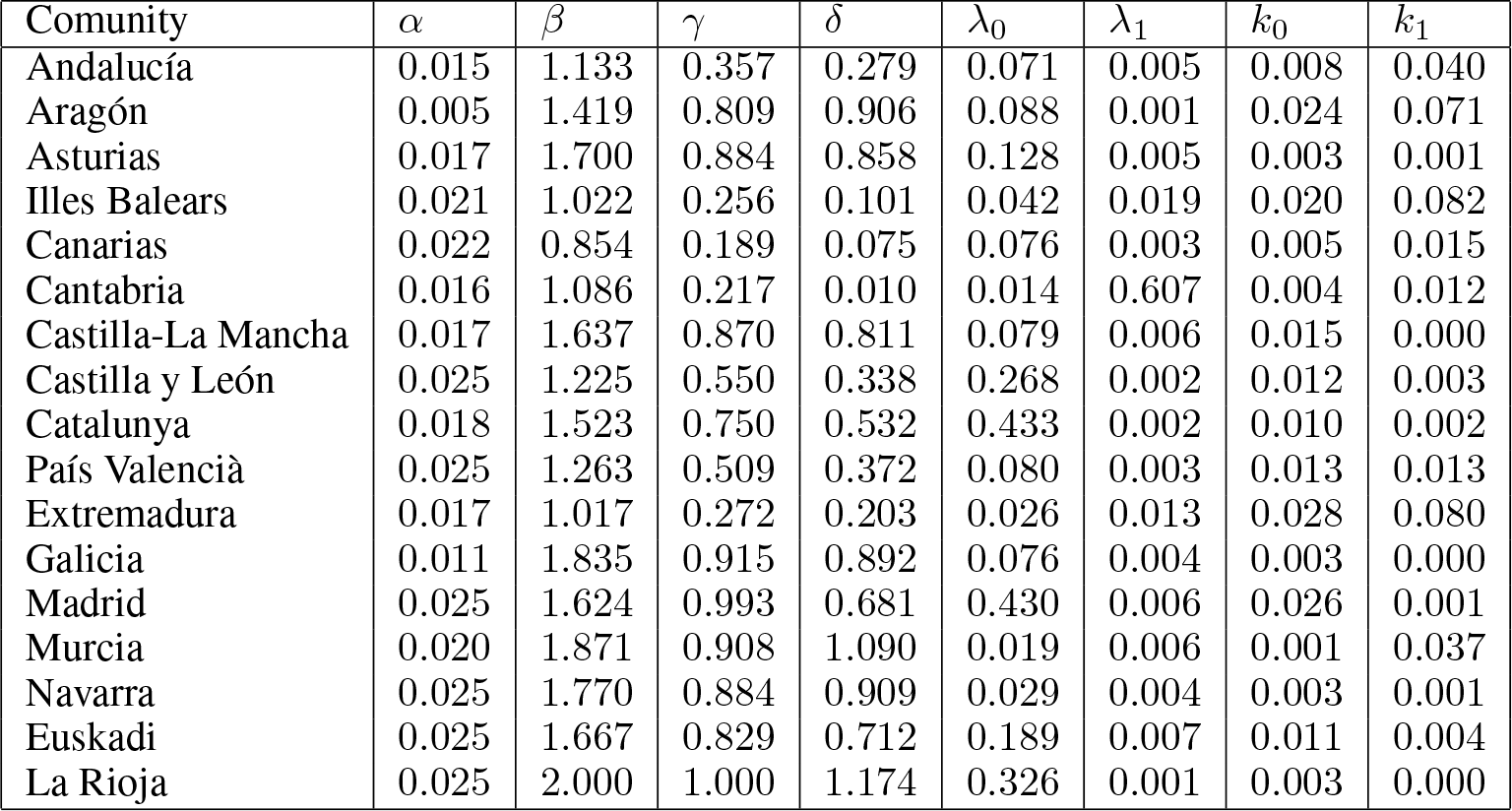
Sample table title

Figure 9 displays model results for the two different scenarios. Bottom panel shows the effect of different measures of confinement since day 1 in an hypothetical control scenario. These dynamics are the result of increasing the fitted *α* until approx 1% daily. This is a significant increase having into account the initial value of protection rate.

**Figure 9:**
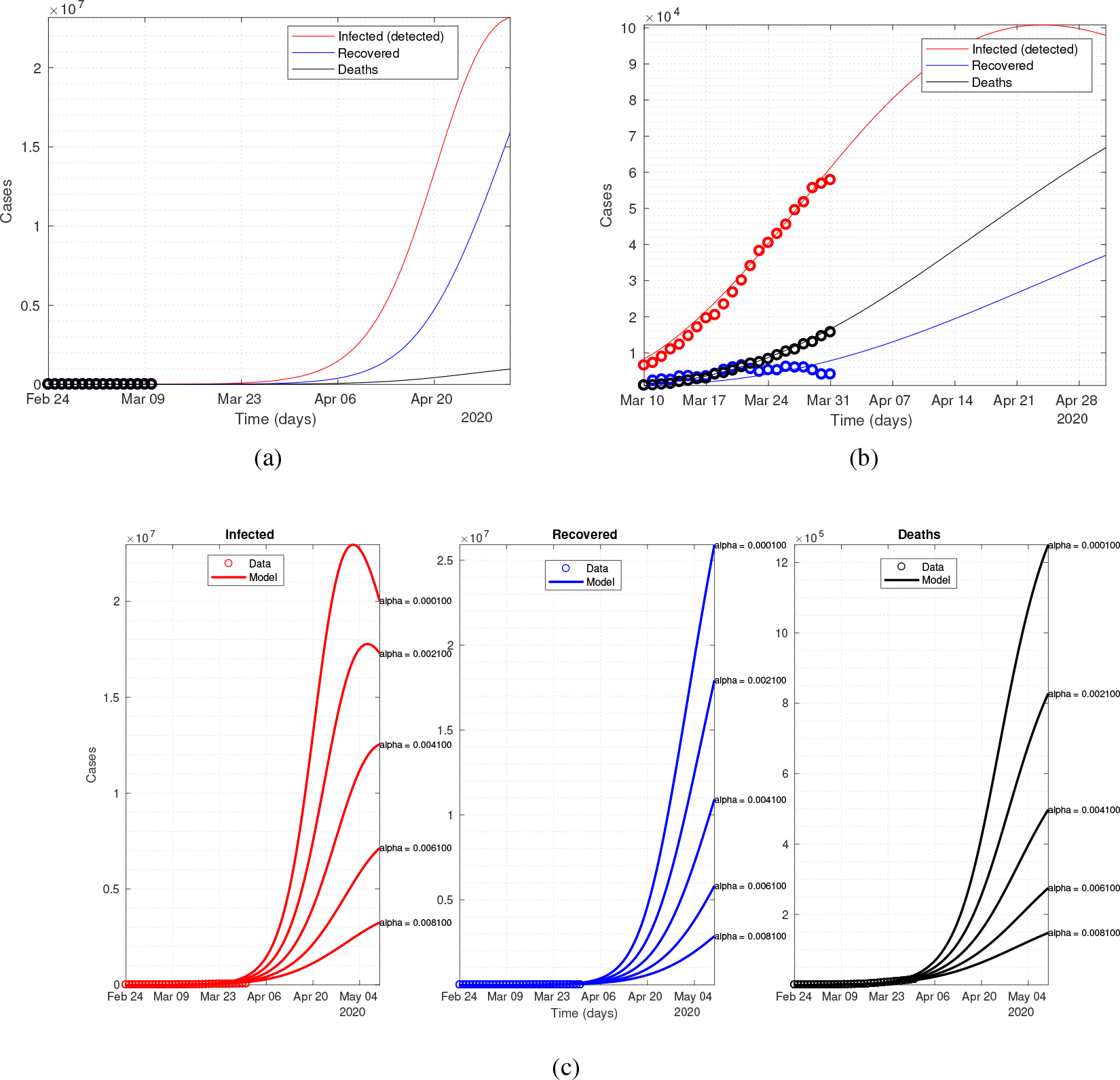
Model fitting and control scenarios for Italy. (a) fitting before confinement restrictions (b)fitting after confinement restrictions and (c) Control scenarios

## 4 Discussion

Current estimates of future trends in new infections in the weeks after March 23 largely compromise the capacity of the Spanish health system, as it happened for Italy. This is especially critical in particular for intensive care units from the end of March [23]. Our projections for the forthcoming evolution in the COVID-19 epidemic curve for Spain show a pessimistic scenario unless current social distancing measures are effective (see Appendix for an update as of March 31. With the estimates at hand, over 1.4 million people would be clinically diagnosed at the disease maximum and over 100.000 resulting deaths. Imposing stricter measures under the current uncertainties seems a logical preventive option given the considerable gains in terms of infected and casualties, even at moderate increases in alpha. Assessment of total country changes by means of aggregated and CCAA disaggregated data estimates, provides similar dynamical evolution despite different absolute values. Discrepancies to larger values might partly come from the heterogeneous population landscape in the different CCAA (e.g. cities with high population density), as opposed to the homogeneous contact assumption in the aggregated model estimates. Gaining extra time for exerting alternative interventions, such as temporary increases in public health responses, or increasing the capacity of massively screening the population in order to gain an accurate value of the real prevalence of SARS-CoV-2, or to develop disease treatment strategies seem a logical conclusion. While two of the most populated CCAA (namely Catalunya with roughly 7.7*M* people and Madrid Community, with 6.7*M* people) are already in the lead in terms of epidemic progression, other similarly populated regions lag only a few days entering a similar slope in their respective epidemic growths. The largest population density in Madrid compared to other CCAA together with its more advanced epidemic progression cast urgent attention on the evolution of this highly active epidemic focus. Those regions where the outbreak was initiated earlier (e.g. Madrid, Catalunya, Euskadi and Navarra) or appear to have more connectivity to the initial foci and largest population density tend to show earlier peaks. Those regions yet on early stages of the exponential increase are those where strong initial social isolation measures (therefore protection of the population), would have largest effects in resulting infected (Figure 2).

The projected rise in the number of infections shown in Figure 1 is very sensitive to the latest data used, as these data are the ones governing the changes in trends, therefore errors in reporting or other large uncertainties associated with these values may dramatically alter the final outcome in terms of number of infected and associated mortality. For the curve to happen as shown, the simulation assumes the initial conditions hold, therefore if that were not the case and measures were lifted before the epidemic reaches *R*_0_*≈*1, infections might rise again. Improvements can be achieved with a more complete integration of time delay coordinates in classical *SEIR* models [6]. This way, both the incubation period and the period before recovery, as well as the precise role of asymptomatic population can be better represented at times when these information is highly uncertain. A substantial gain in the epidemic projection in the form of reduced infections in the population would already be achieved with a four-times rise in the efficacy of the control situation we had around March 9*−*16. This was the time when social distancing/confinement was imposed (March 13). Time will show to which extent current measures manage to increase the value of alpha. Application of strict containment measures clearly show drastic effect on the epidemic progression and a substantial reduction in both infected and to a lesser extent, deaths (Figure 5).

The control scenarios presented contemplate a significant increase in the percentage of the population under confinement, ranging from 1.3 percent daily to 13 percent daily. These percentages imply that after a week of starting this control, the percentage of protected susceptible population is 9.1% and 91%, respectively, from the start of the measurement. These numbers explain the drastic differences in the two dynamics observed and the huge gain in lowering the toll of infected.

Update of the epidemic situation in Spain partially showing the effect of partial lockdown and the current values for alpha are shown in the Appendix as for March 31.

## 5 Conclusions

The Covid-19 pandemic is exerting an unprecedented stress on the public health systems of many countries. Those at major risk now are Italy and Spain, and for them, efficacy of the partial confinement or total lockdown effects are yet unknown. Under this situation, we implemented a modified SEIR compartmental model accounting for infection from undiagnosed individuals and for different levels of population isolation, to evaluate effects of contacts reduction in the epidemic temporal dynamics. Among the advantages of the implemented model, it should be noted that despite the simplicity of the hypotheses, the adjustments obtained were accurate and the projections made do not differ much from those other of more complex models. Also instantaneous increment of cumulative diagnosed people depends on the history of cumulative infected people, by which the latent period can be taken into consideration. The results of the implemented control scenarios for March 23 show that a drastic isolation of the susceptible population should be implemented as soon as possible. For even not so drastic increases in alpha (two or three times the current rate) imply also significant reductions in the incidence of cases. The adjustments made in different CCAA also serve to verify the efficacy of the isolation hypothesis for the most affected communities (Madrid and Catalunya). They also serve as a basis for timely action in those communities that do not yet have a significant number of cases that jeopardises their health systems. Policymakers should weigh in the values and ethical considerations of employing now maximum strength in actions to help reduce the slope of the epidemic curve against the enormous associated economic cost. However, our study indicates that a three-weeks interruption of labor activities, thereby a drastic reduction in contacts, could end the current epidemic in around two months and drastically reduce both the burden of this disease, and much lower the toll of lives. Our results could also provide useful suggestions for the prevention and control of the COVID-19 outbreaks in different countries and locations such as Argentina and USA lagging the current epidemic wave in Spain and Italy. These represents two very different scenarios. In the case of the USA, no radical control measure was initially implemented by the federal government and the epidemic seems to be in a phase of uncontrolled growth. On the Argentina side, the health authorities seem to have taken note of what is happening in Europe and more strict movement restrictions have been implemented, although it is difficult to determine the degree of commitment by the population.

However, our model does not consider space explicitly as we approached aggregated data. This lack of spatial granularity may affect the accuracy of simulations when the spread of infections in a territory reaches and takes off in a populated city. The former might alter results producing more new cases than expected, exactly what the aggregation for CCAA data clearly shows. Spatial modelling should also be incorporated explicitly and extension of this modelling approach to incorporate movement of individuals should follow. Results should be interpreted with care as projections at these initial stages of the epidemic are very dependent on the quality of data, with small changes in observed values producing large variations in trends. However, even with this limitation in mind, the magnitude of positive increments in cases suggests these results are strong. Variable isolation strength measures can be tested with this model and inform governments of the most probable effects of their actions on the initial course of the COVID-19 disease.

## Data Availability

The data come from open data sources that are linked bellow.

https://www.mscbs.gob.es/profesionales/saludPublica/ccayes/alertasActual/nCov-China/home.htm

https://experience.arcgis.com/experience/50d6c4c5599a43f4bebf517daa3a97b6

https://grafana.sysadm.es/d/Y9Anj9_Wz2dsadasDXCAxz5/coronavirus-spain?orgId=4&refresh=1h

## A Updated Results for Spain

In this appendix we show an update of the fitted model with the data reported up to March 31. Figure 10 shows the the fitting and projections of the model with the new data. If we compare these results with those in the previous sections, the model now predicts an epidemic peak much lower than before. This may be mainly because of the Spanish government control measures imposed and a harder confinement restriction. The model parameters now are *α* = 0.087, *β* = 1.305, *γ* = 1.044, *δ* = 0.510, *λ* = [0.095, 0.017], *k* = [0.026, 0.022]. A significant increase in the control rate is mostly evident.

**Figure 10:**
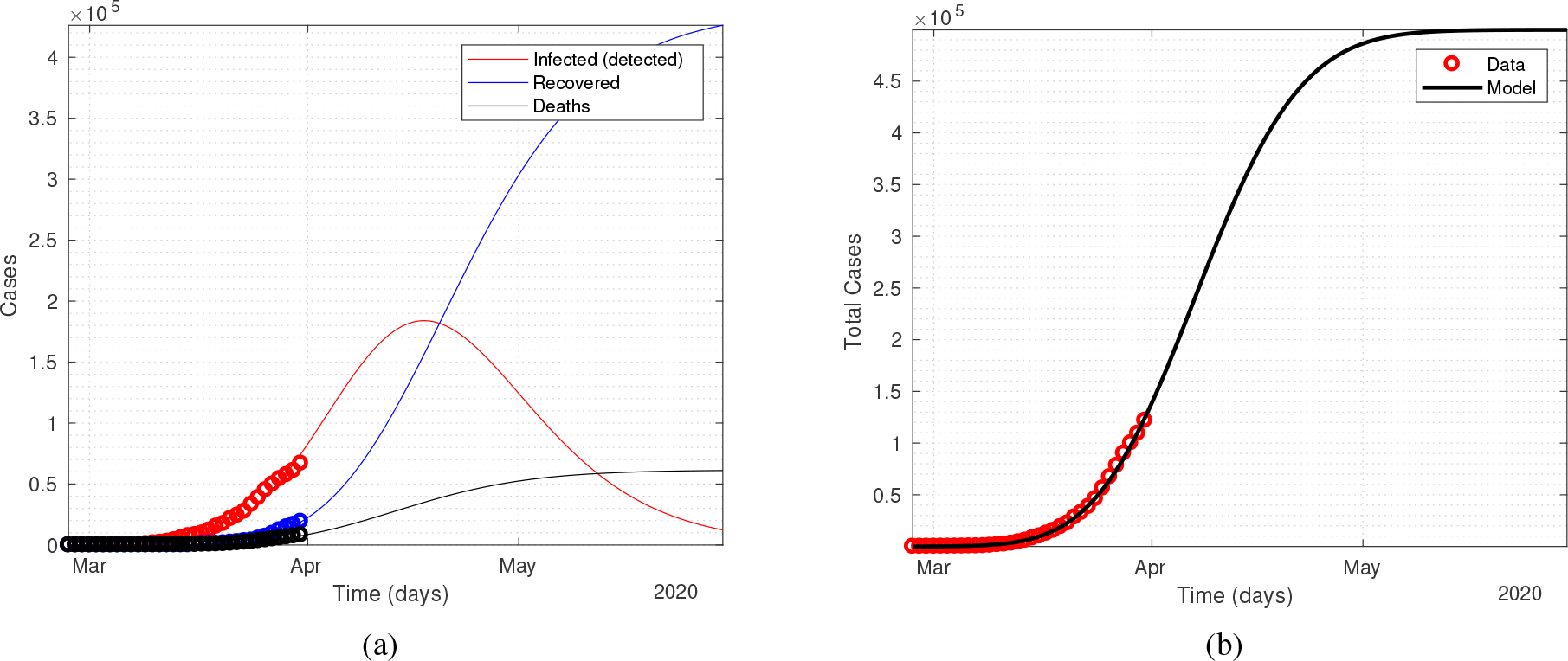
Model fitting for Spain updated to March 31. (a) total active cases, recovered and deaths (b) total confirmed cases.

In Figure 11 and to better understand the current scenario in the context of the scenarios previously outlined in Figure 5, the dynamics corresponding to the parameters indicated above are added as dashed lines for each panel (Reported, Recovered and Dead).

**Figure 11:**
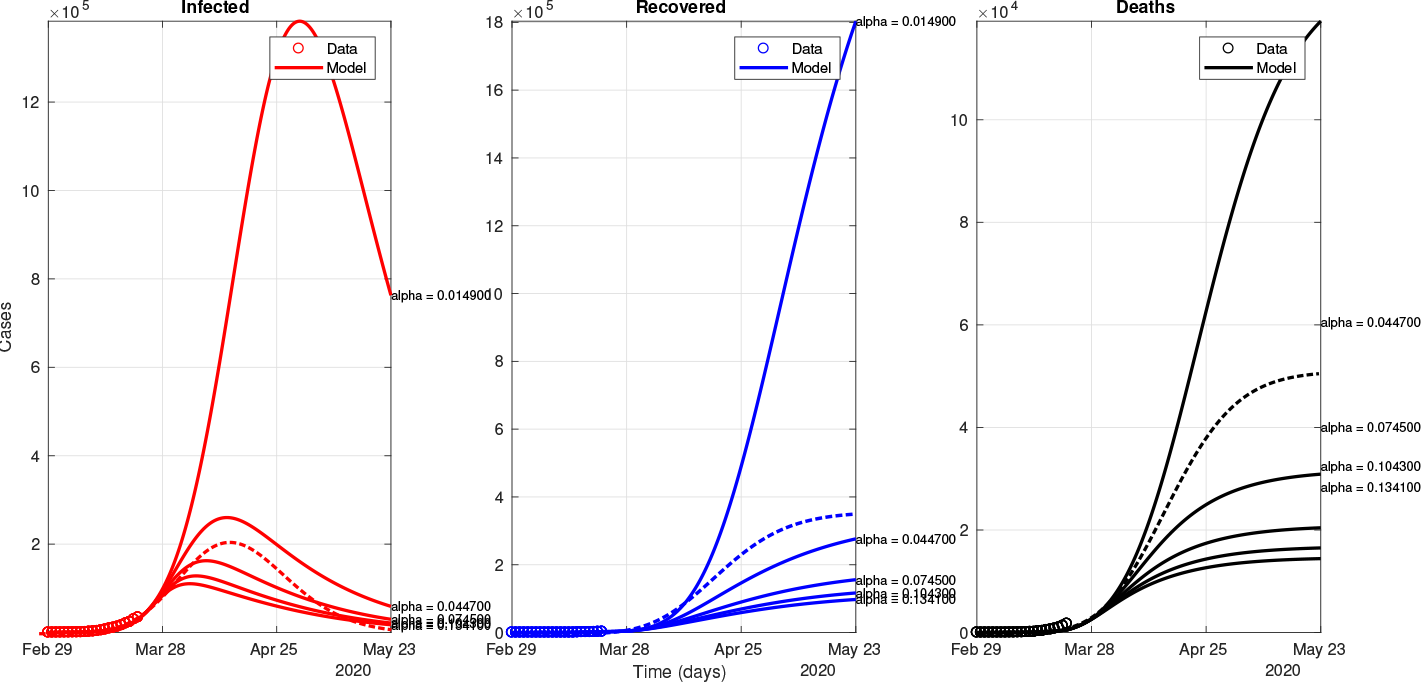
Control scenarios for the global model (all Spain) with the predictions as for March 31st added as dashed lines.

## B Updated scenario for each CCAA in Spain

In the same way than we did for Spain, in this section we show results of the model fitting with the data updated until March 31*st*. In Figure 12 the new fittings are showed and in Table 3 the fitted parameters are listed. Notice the closeness to the epidemic peak in some of the CCAA, a fact that can be real or be due to the yet low prevalence of the disease in parts of Spain.

**Table 3:**
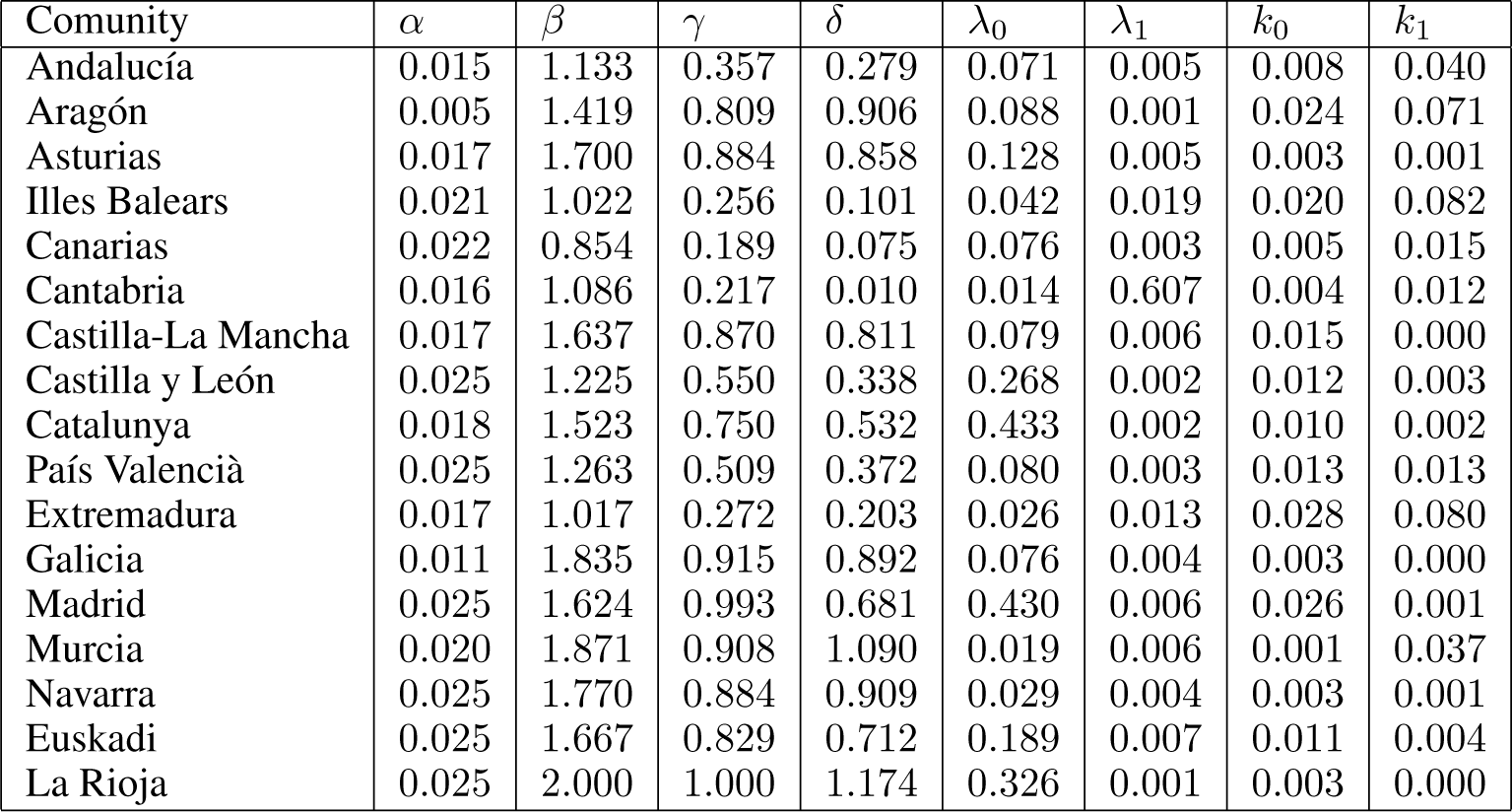
Sample table title

**Figure 12:**
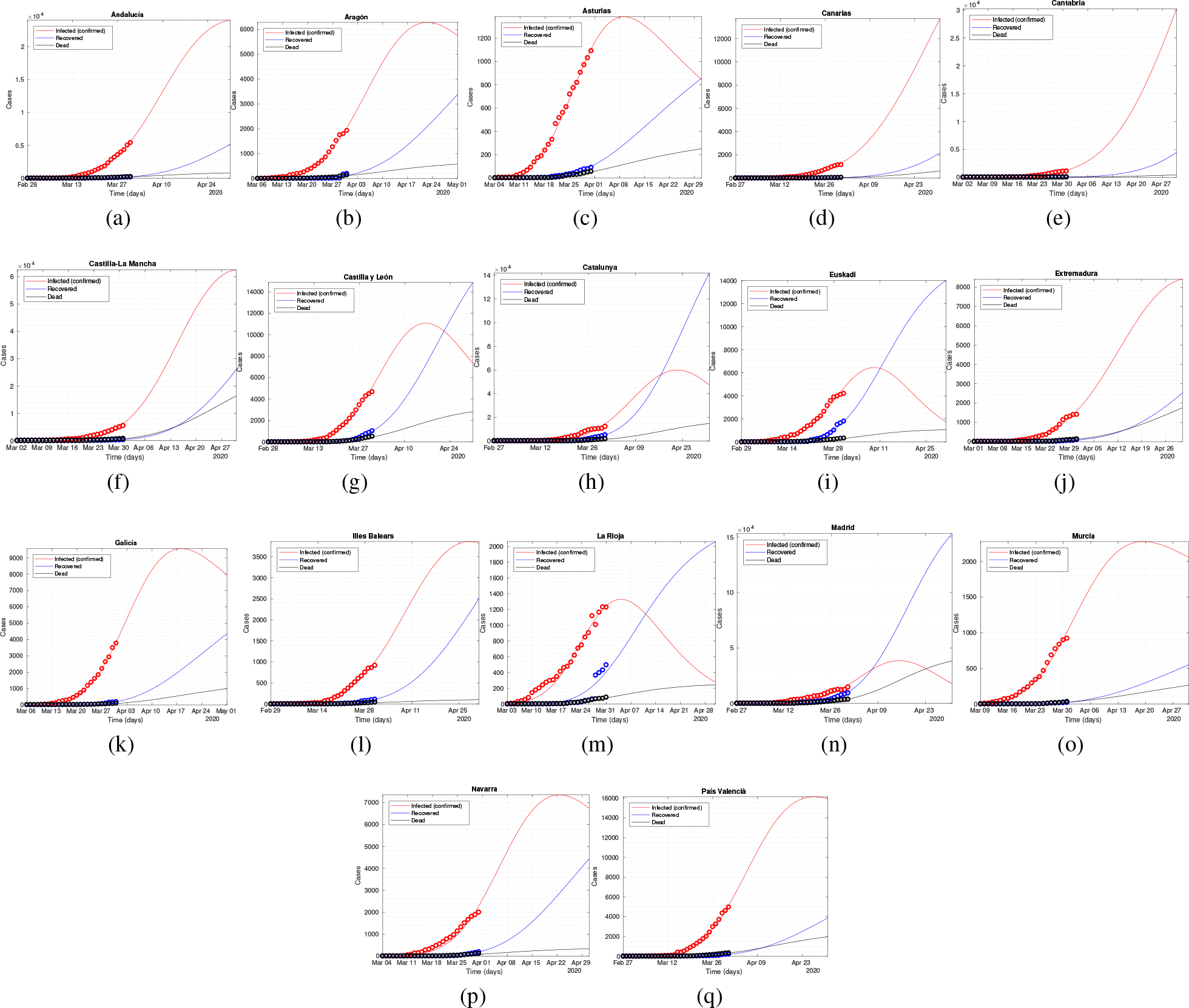
Model extended fitting for the 17 CCAA of Spain.

## C Increasing the actual protection rate for Italy

With the purpose of making a new evaluation of what an increase in the current protection rate for Italy might represent, several control scenarios were run where the alpha value was gradually increasing from 3.5% daily up to the post-control scenario assumption of 7.2% daily. Results can be seen in Figure 13.

**Figure 13:**
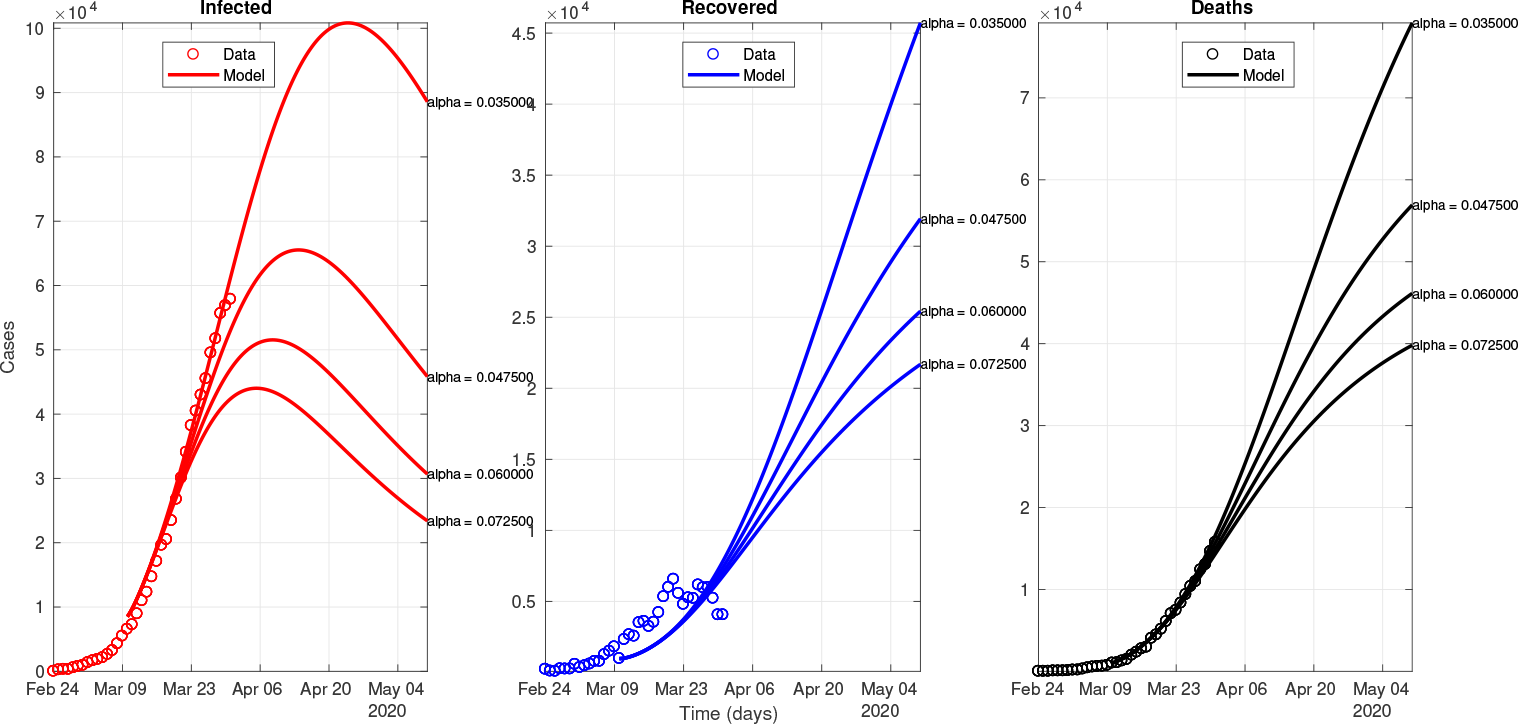
Control scenarios for Italy using as a starting point the parameters obtained with the post-control fitting (Table2)

